# Projected Impact of Nonpharmacologic Management of Stage 1 Hypertension Among Lower-Risk US Adults

**DOI:** 10.1101/2023.12.26.23300563

**Authors:** Kendra D. Sims, Pengxiao Carol Wei, Joanne M. Penko, Susan Hennessy, Pamela G. Coxson, Nita H. Mukand, Brandon K. Bellows, Dhruv S. Kazi, Yiyi Zhang, Ross Boylan, Andrew E. Moran, Kirsten Bibbins-Domingo

## Abstract

**Background:** The 2017 American College of Cardiology (ACC)/American Heart Association (AHA) guidelines newly classified 31 million US adults as having stage 1 hypertension. The ACC/AHA guidelines recommend behavioral change without pharmacology for the low-risk portion of this group. However, the nationwide reduction in cardiovascular disease (CVD) and associated healthcare expenditures achievable by evidence-based dietary improvements, sustained weight loss, adequate physical activity, and alcohol moderation remain unquantified. We estimated the effect of systolic BP (SBP) control and behavioral changes on 10-year CVD outcomes and costs.

**Methods:** We used the CVD Policy Model to simulate CVD events, mortality, and healthcare costs among US adults aged 35-64. We simulated interventions on a target population, identified from the 2015-2018 National Health and Nutrition Examination Survey, with low-risk stage 1 systolic hypertension: defined as untreated SBP 130-139 mmHg and diastolic BP <90 mmHg; no history of CVD, diabetes, or chronic kidney disease; and low 10-year risk of CVD. We used published meta-analyses and trials to estimate the effects of behavior modification on SBP. We assessed the extent to which intermittent healthcare utilization or partial uptake of nonpharmacologic therapy would decrease CVD events prevented.

**Results:** Controlling SBP to <130 mmHg among the estimated 8.8 million U.S. adults (51% women) in the target population could prevent 26,100 CVD events, avoid 2,900 deaths, and save $1.6 billion in healthcare costs over 10 years. The Dietary Approaches to Stop Hypertension (DASH) diet could prevent 16,000 CVD events among men and 12,000 among women over a decade. Other nonpharmacologic interventions could avert between 3,700 and 19,500 CVD events. However, only 5.5 million (61%) of the target population regularly utilized healthcare where recommended clinician counseling could occur.

**Conclusions:** As only two-thirds of U.S. adults with Stage 1 hypertension regularly receive medical care, substantial benefits to cardiovascular health and associated costs may only stem from policies that promote widespread adoption and sustained adherence of nonpharmacologic therapy. Future work should quantify the population-level costs, benefits, and efficacy of improving the food system and local infrastructure on health behavior change.

**Clinical Perspective:** **What is new?**

- Guidelines recommend modifying health behaviors to achieve blood pressure control in individuals with stage 1 hypertension, but the nationwide reduction in cardiovascular disease (CVD) and associated healthcare expenditures achieved remain unquantified.
- Using a public policy simulation model of CVD, we projected that controlling stage 1 systolic hypertension with nonpharmacologic therapy among an initially low-risk population of nearly 9 million young- and middle-aged adults would avert approximately 26,000 CVD events, 3000 deaths, and $1.6 billion in healthcare costs over 10 years.
- The Dietary Approaches to Stop Hypertension (DASH) diet could provide the greatest population benefit.

**What are the clinical implications?**

- Evidence-based health behaviors, such as adopting the unprocessed foods-centric Dietary Approaches to Stop Hypertension (DASH) diet, could provide long-term dividends to improved cardiovascular health.
- However, one-third of initially low-risk adults with stage 1 hypertension did not regularly use healthcare. There additionally are documented challenges to sustaining these lifestyle changes. Systemic deprivation of health-promoting resources in the social and built environment can pose insurmountable economic barriers for marginalized patients, perpetuating cardiovascular disparities.
- The recommended medical provider counseling on behavioral modification must be paired with community interventions, infrastructure improvements, and nutrition-promoting food system policies to promote adherence.

## Introduction

The 2017 American College of Cardiology and American Heart Association (ACC/AHA) guidelines reinforced the importance of blood pressure (BP) control for disease prevention by lowering the hypertensive threshold^1^. Compared with the Seventh Report of the Joint National Committee on Prevention, Detection, Evaluation, and Treatment of High Blood Pressure (JNC7) BP classification, the ACC/AHA guidelines newly categorizes 31 million US adults as having stage 1 hypertension (systolic BP [SBP]/diastolic BP [DBP] 130/80-139/89 mmHg) ^2,3^. However, the guidelines recommend pharmacotherapy only for the small subset of this group who are 65 years and older, have a history of cardiovascular disease (CVD), diabetes, chronic kidney disease, or a 10-year risk of CVD of at least 10 percent. For the rest of this patient base, clinicians are instead advised to provide counseling on lifestyle changes based on findings from short-term behavioral intervention trials^4^. These nonpharmacologic therapies include the Dietary Approaches to Stop Hypertension (DASH) diet^5^, sufficient physical activity^6^, balancing dietary intake of potassium to sodium^7^, sustaining loss of excess body weight^8^, and ceasing excessive alcohol consumption^9^. The potential long-term population health and economic benefits of scaling up behavioral intervention trial findings for the population with stage 1 hypertension not recommended pharmacotherapy remain unquantified^10^. Additionally, the extent to which lack of regular healthcare access might limit opportunities to identify stage 1 hypertension and counsel nonpharmacologic therapies that prevent CVD has not been ascertained for this group.

Using a public policy simulation model of CVD and data from the National Health and Nutrition Examination Survey (NHANES), we first described the US adult population with stage 1 systolic hypertension (SBP 130-139 mmHg and DBP <90 mmHg) who would be recommended only nonpharmacologic therapy by the 2017 ACC/AHA BP guideline. We then projected the effect of SBP control via nonpharmacological therapy on CVD incidence, CVD mortality, and healthcare costs over 10 years. We also assessed the attenuated impact of recommended medical provider counseling on nonpharmacologic therapy in this population due to lack of regular healthcare utilization.

## Methods

### Model overview

The CVD Policy Model is a state-transition computer simulation model of coronary heart disease (CHD) and stroke incidence, recurrence, mortality, and direct healthcare costs in all US adults aged 35-94 years^11–15^. Core model inputs are estimated from nationally representative databases, longitudinal cohort studies, and natural history studies (Supplemental Table S2). For the population without CVD, the model predicts the annual incidence of CHD, stroke, and non-CVD mortality via competing risk models. We base estimates on age, sex, and CVD risk factors, using risk functions calculated from data pooled and harmonized from several National Heart, Lung, and Blood Institute (NHLBI) observational cohorts^16–20^. The population distribution of CVD risk factors (i.e., SBP, DBP, self-reported use of antihypertensive medications, low- and high-density lipoprotein cholesterol, exposure to tobacco smoke, and diabetes status) are derived from the NHANES 2015-2016 and 2017-2018 cycles^21^. By incorporating expected population growth as well as changes in risk factors and event rate, the model projects subsequent CVD events, coronary revascularization procedures, and mortality from CVD and non-CVD causes in annual cycles (Supplemental Figure S1). Direct healthcare costs associated with acute CVD events are derived from the National Inpatient Sample (NIS) and costs associated with chronic CVD and background (non-CVD) healthcare are derived from the Medical Expenditure Panel Survey (MEPS)^22,23^. The model was calibrated to reproduce annual CVD event rates within 1% of observed hospital discharges in NIS (2012-2016) and deaths observed in US National Vital Statistics System (2018)^22,24,25^. The study was considered exempt by the Institutional Review Board at the University of California, San Francisco. We followed the Consolidated Health Economic Evaluation Reporting Standards (CHEERS) reporting guideline for health economic evaluations^26^.

### Model inputs and target population

Key model inputs are described in Supplemental Table S2. The target population is US adults aged 35-64 years with untreated stage 1 systolic hypertension (SBP 130-139 mmHg and DBP <90 mmHg) for whom only nonpharmacological therapy (e.g., dietary changes, increasing physical activity, weight loss) is recommended by the 2017 ACC/AHA BP guideline^1^. We limited our study to stage 1 systolic hypertension due to insufficient evidence for nonpharmacologic therapy on DBP on CVD outcomes or long-term CVD outcomes associated with stage 1 isolated diastolic hypertension (SBP <130 mmHg and DBP 80-89 mmHg) ^27^. The mean of all available measures for each NHANES participant was used for BP. Operationally, we used the subsample of NHANES of appropriate age and blood pressure ranges; no reported current medications to treat high BP; no history of CVD, diabetes, or chronic kidney disease; and the contemporaneous criteria for <10% predicted risk of a CVD event in 10 years^28^ (Supplemental Table S3).

Based on current US guidelines for physical activity^29^ and diet^30^, we estimated the proportion of men and women in the target population who are insufficiently active (<600 metabolic equivalent minutes of task [METS] per week), overweight or obese (body mass index [BMI] ≥25 kg/m^2^), and who consume excessive sodium (≥2300 mg/day), inadequate potassium (<3400 mg/day for men; <2600 mg/day for women), lower than recommended amounts of fruits and vegetables (<2 cups of fruit and/or <2.5 cups of vegetables per day), and excessive alcohol (>2 drinks/day for men; >1 drink/day for women; 14 grams of alcohol = 1 drink). We assumed that no one in the target population adhered to the DASH diet based on low reported intake of fruits and vegetables and excessive sodium in NHANES. We implemented a classification of regular access to healthcare previously used in NHANES^31^, based on participants confirming having some form of health insurance, at least one routine place they receive healthcare, and at least one visit to a healthcare professional in a non-emergency room context in the past year.

### Simulated comparators

We simulated a baseline “status quo” scenario that incorporated SBP changes expected with demographic shifts as the target population ages. We compared the status quo with the mean SBP reduction from trial and meta-analysis data of nonpharmacological management among individuals in the target population not meeting recommended thresholds for specific health behaviors (Supplemental Table S1). Adopting the DASH dietary pattern reduced SBP by 5.5 mmHg ^32^. Sustained weight loss among adults with a BMI ≥25 kg/m^2^ reduced SBP by 4.8 mmHg ^33^. Reduced dietary sodium among adults with excessive intake reduced SBP by 4.2 mmHg ^34^. Increased fruit and vegetable consumption among adults with insufficient dietary potassium reduced SBP by 2.8 mmHg ^32^. Adequate physical activity among adults who do not engage in sufficient weekly METS reduced SBP by 3.8 mmHg ^35^. Reduced alcohol consumption among adults consuming more than recommended drinks weekly reduced SBP by 3.1 mmHg ^36^. The total population effect of an intervention was therefore dependent both on the size of the SBP reduction and number of individuals exhibiting the modifiable risk. Since the mean SBP in the target population overall is less than 4 mmHg above 130 for both men and women, some of these effect sizes reduced blood pressure below the 129 SBP target assumed in the initial analysis. As trial and meta-analysis data of nonpharmacological management did not quantify either selection into their studies or long-term adherence, we simulated in sensitivity analyses control to 129 SBP assuming 50% of the intervenable target population adhered to each intervention over 10 years.

We also compared the “status quo” scenario with a scenario controlling SBP to <130 mmHg (i.e., max SBP set to 129 mmHg) each year for all individuals in the target population. We conservatively assumed that those whose risk factor profile changed so that they would be eligible for pharmacological intervention according to the 2017 ACC/AHA BP guideline (e.g., developed diabetes) would initiate medications, resulting in an SBP <130 mmHg with no change to their other risk factors. We also simulated controlled SBP <130 mmHg only among those with regular access to healthcare. This set of analyses assessed the full benefit of the entire target population achieving the goals outlined by the 2017 ACC/AHA guidelines, as well as how focusing on counseling in the healthcare setting would attenuate the extent of CVD events prevented and healthcare costs saved.

### Statistical analyses

We simulated a nationally representative dynamic cohort of US adults over 10 years, starting in 2018 and running through 2027. For each intervention, we calculated the difference in (1) incident and total CVD events, (2) CVD mortality, and (3) associated direct healthcare costs relative to the status quo. Using Monte Carlo methods, we repeated the analyses 1000 times, randomly selecting model input parameters with replacement from pre-specified distributions scaled to the mean and confidence interval (Supplemental Table S3) and generated 95% uncertainty intervals around model estimates. We used normal distributions for baseline SBP and mean SBP changes expected for nonpharmacologic therapy ^21,32–36;^ lognormal distributions for the relative risks of CHD, ischemic stroke, and hemorrhagic stroke associated with SBP changes ^37^; and lognormal distributions for healthcare utilization costs ^22,23^.

The CVD Policy Model is programmed using the Intel Fortran Compiler Classic 2021.5 and Monte Carlo simulations are run using Python version 3.10. We estimated model inputs using Stata 17 and SAS version 9.4 and analyzed outcomes using Microsoft Excel version 2108.

## Results

The simulated target population represents an estimated 4.2 million men and 4.6 million women aged 35-64 years. Table 1 presents the gender-stratified clinical and behavioral characteristics. Only 40% reported adequate intake of fruits and vegetables. Four-fifths had a BMI classified as either overweight or obese. Men were more likely than women to consume excess sodium (92% versus 71%) and alcohol (28% versus 17%). Insufficient physical activity was more prevalent among women than men (45% versus 30%). Only 51% of men and 73% of women in the target population were both insured and had visited their routine, non-emergency healthcare provider within the past year.

**Table 1.** Characteristics of U.S. population aged 35-64 years with 2017 ACC/AHA stage 1 systolic hypertension recommended nonpharmacologic therapy,* NHANES years 2015-2018.

| Characteristics | Stage 1 systolic hypertension<sup>†</sup> |  |
| --- | --- | --- |
|  | Men<br>(N=152) | Women<br>(N=186) |
| Age, years | 47.0 (1.4) | 50.4 (0.7) |
| Systolic BP, mmHg | 133.6 (0.3) | 133.7 (0.3) |
| Diastolic BP, mmHg | 78.5 (0.9) | 76.5 (0.9) |
| Predicted 10-year CVD risk, % | 4.7 (0.4) | 2.3 (0.2) |
| **Non-pharmacological risk factors** |  |  |
| Overweight or obese, % | 80.2 | 78.6 |
| Physically inactive, % | 29.9 | 44.8 |
| High sodium intake, % | 91.9 | 71.1 |
| Low fruit and vegetable intake, % | 57.4 | 62.6 |
| High alcohol consumption, % | 27.9 | 16.6 |
| **Healthcare access** |  |  |
| Health insurance type, % |  |  |
| Public insurance | 6.2 | 11.3 |
| Private insurance | 77.8 | 78.5 |
| No insurance | 16.0 | 10.2 |
| Regular source of healthcare, % | 71.3 | 82.9 |
| Healthcare visits in past year, % |  |  |
| None | 32.5 | 11.6 |
| One | 19.4 | 28.3 |
| Two or more | 48.1 | 60.1 |
| Values are mean (standard error) or %. Estimates calculated using survey sample weighting procedures on data collected from National Health and Nutrition Examination Survey years 2015-2018. N reflects the number of NHANES participants for each subgroup.<br>ACC = American College of Cardiology; AHA = American Heart Association; BP = blood pressure; NHANES – National Health and Nutrition Survey<br>*The 2017 ACC/AHA guideline defines stage 1 hypertension as having a systolic blood pressure of 130 to 139 mmHg or a diastolic blood pressure of 80 to 89 mmHg and recommends non-pharmacological BP management for those without diabetes, chronic kidney disease, or cardiovascular disease and less than 10% risk of a CVD event within 10 years calculated using the Pooled Cohort Risk Equations<br><sup>†</sup> Stage 1 systolic hypertension defined as systolic BP ranging from 130 – 139 mmHg and diastolic BP < 90 mmHg |  |  |

By focusing on stage 1 systolic hypertension, we target approximately half of the total population for whom the ACC/AHA 2017 guidelines recommend behavioral change. These guidelines would classify an additional 3.9 million men and 3.2 million women at low-CVD risk as having stage 1 isolated diastolic hypertension (Supplemental Table S4).

Compared with the status quo among US adults with stage 1 hypertension recommended nonpharmacologic therapy, controlling SBP to 129 mmHg is projected to prevent 26,100 (95% CI: 22,600 – 30,700) incident CVD events and 2,900 (95% CI: 2,400 – 3,300) CVD deaths over 10 years (Table 2). Averting these CVD events and mortality would save an estimated $1.6 (95% CI: $1.4 – $1.9) billion in hospitalizations, procedures, outpatient care, and background healthcare costs. Figure 1 displays how not intervening among men and women who did not regularly utilize healthcare would reduce incident CVD events prevented by each evidence-based intervention by 39%. By intervening only via provider counseling, the projected number of CVD events and CVD deaths prevented by controlling SBP to <130 mmHg was reduced, respectively, to 15,800 (95% CI: 13,800 – 18,600) and 1,800 (95% CI: 1,600 – 2,000).

**Figure 1.**
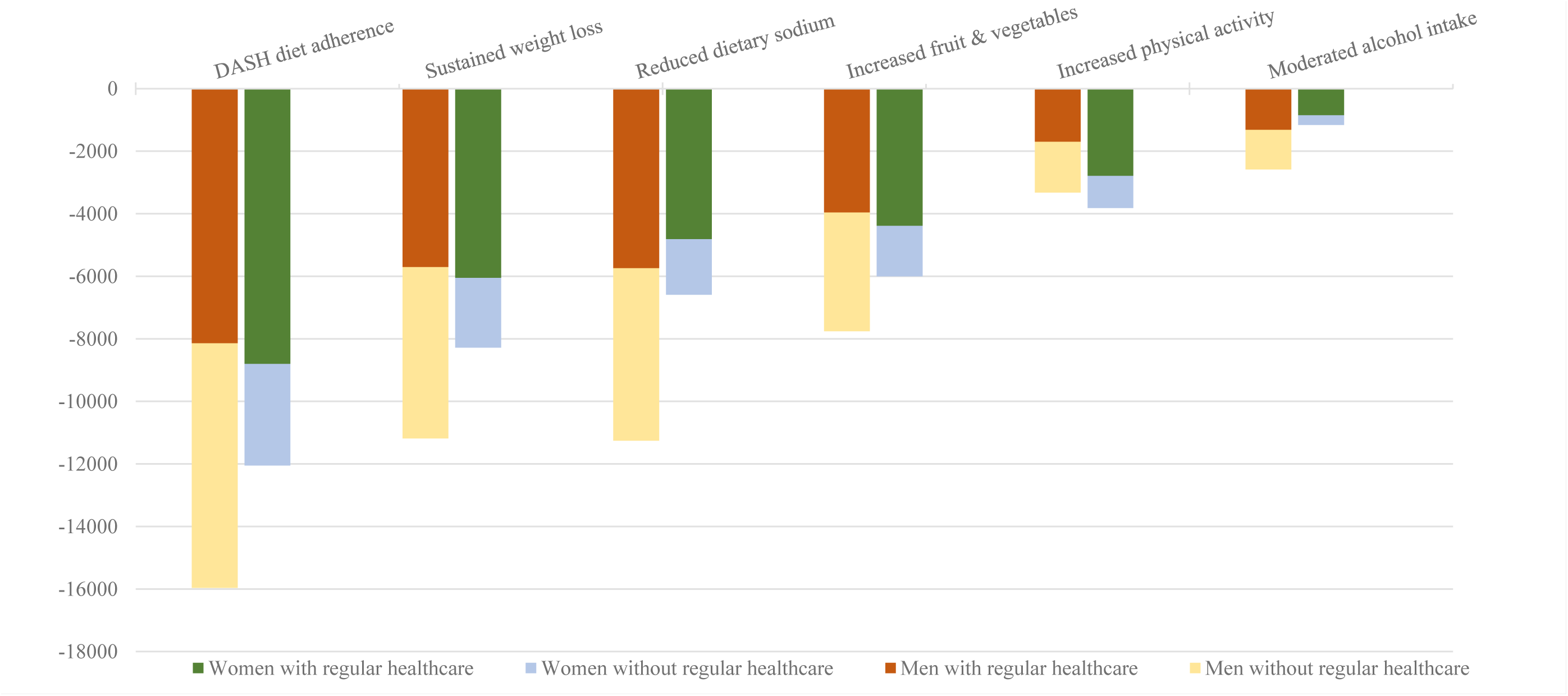
Projected 10-year reduction in incident cardiovascular disease events for evidence-based non-pharmacological approaches to reducing blood pressure among adults with and without self-reported regular healthcare utilization. Simulations targeted adults with stage 1 systolic hypertension (SBP 130 – 139 mmHg and DBP <90) who qualify for nonpharmacological therapy in the 2017 ACC/AHA guidelines. We simulated effect of the DASH diet in those not currently eating a DASH dietary pattern (assumed to be everyone in the target population), sustained weight loss in adults who are overweight or obese (80% of men and 79% of women), reduced sodium intake in those currently consuming ≥ 2300 mg/day (92% of men and 71% of women), increasing fruit and vegetable intake in those not currently eating at least 2.5 cups of vegetables and 2 cups of fruit per day (94% of men and 96% of women), sufficient physical activity for inactive adults (<600 weekly metabolic physical equivalents of task; 30% of men and 45% of women), and reductions in alcohol intake for men and women drinking more than 2 or 1 drink per day, respectively (28% of men and 17% of women). 49% of men and 27% of women reported not having regular healthcare: i.e., being uninsured, not having a routine place to receive healthcare, or not visiting a healthcare professional in a non-emergency room context in the previous 12 months.

**Table 2.** Projected impact of controlling SBP to under 130 mmHg in adults with stage 1 hypertension recommended only nonpharmacologic therapy by the 2017 ACC/AHA BP guideline*, 2018-2027.

| Number of events prevented and healthcare costs saved (95% Confidence Intervals) |  |  |  |  |  |  |
| --- | --- | --- | --- | --- | --- | --- |
| Simulated target group | Population reached (millions) | Incident CVD** | Total myocardial infarctions*** | Total strokes**** | CVD mortality | Healthcare cost savings ¥ (millions 2019 USD) |
| Men |  |  |  |  |  |  |
| Full target population | 4.2 | 15,300<br>(13,100 – 18,200) | 4,100<br>(3,300 – 5,000) | 5,400<br>(4,500 – 6,600) | 1,600<br>(1,300 – 1,800) | \$955<br>(\$832 – \$1,131) |
| Subgroup with regular healthcare access | 2.1 | 7,700<br>(6,600 – 9,200) | 2,100<br>(1,700 – 2,500) | 2,800<br>(2,300 – 3,400) | 800<br>(700 – 900) | \$486<br>(\$423 – \$575) |
| Women |  |  |  |  |  |  |
| Full target population | 4.6 | 10,800<br>(9,500 – 12,500) | 2,600<br>(2,200 – 3,100) | 5,000<br>(4,200 – 6,000) | 1,300<br>(1,100 – 1,500) | \$712<br>(\$624 – \$830) |
| Subgroup with regular healthcare access | 3.4 | 8,100<br>(7,200 – 9,400) | 2,000<br>(1,700 – 2,300) | 3,800<br>(3,200 – 4,500) | 1,000<br>(900 – 1,100) | \$534<br>(\$468 – \$621) |

Values are numbers of events and healthcare costs when controlling systolic BP to 129 mmHg compared to status quo systolic BP over 10 years.
ACC = American College of Cardiology; AHA = American Heart Association; CVD = cardiovascular disease
\*Simulations include those with systolic BP ranging from 130 to 139 mmHg, diastolic BP under 90 mmHg and no indicator of high CVD risk (diabetes, chronic kidney disease, cardiovascular disease, or $\geq 10\%$ predicted risk of a CVD event within 10 years)
\*\*Includes myocardial infarction, cardiac arrest, angina, ischemic stroke and hemorrhagic stroke occurring in the CVD free population
\*\*\*Includes myocardial infarction or ischemic stroke and hemorrhagic stroke occurring in the population with and without history of CVD
¥ Total healthcare costs are undiscounted and include costs from hospitalizations for MI, stroke, cardiac arrest, and revascularization procedures; chronic CVD outpatient care; and background costs for non-cardiovascular care

Full adoption of the DASH diet is the nonpharmacologic therapy that would translate to the greatest mean SBP reduction, preventing a projected 28,000 (95% CI: 18,100 – 38,400) CVD events and saving $1.8 (95% CI: $1.2 – $2.4) billion in healthcare costs (Table 3, Figure 1). Implementing an intervention that sustained weight loss would prevent 19,500 (95% CI: 15,200 – 24,200) CVD events over 10 years. The other nonpharmacologic therapies are projected prevent thousands of CVD events and mortality, with hundreds of millions of dollars in healthcare savings. We quantified in Supplemental Figure S2 the magnitude to which health and economic benefit would be drastically attenuated by only achieving 50% adherence of each intervention.

**Table 3.**
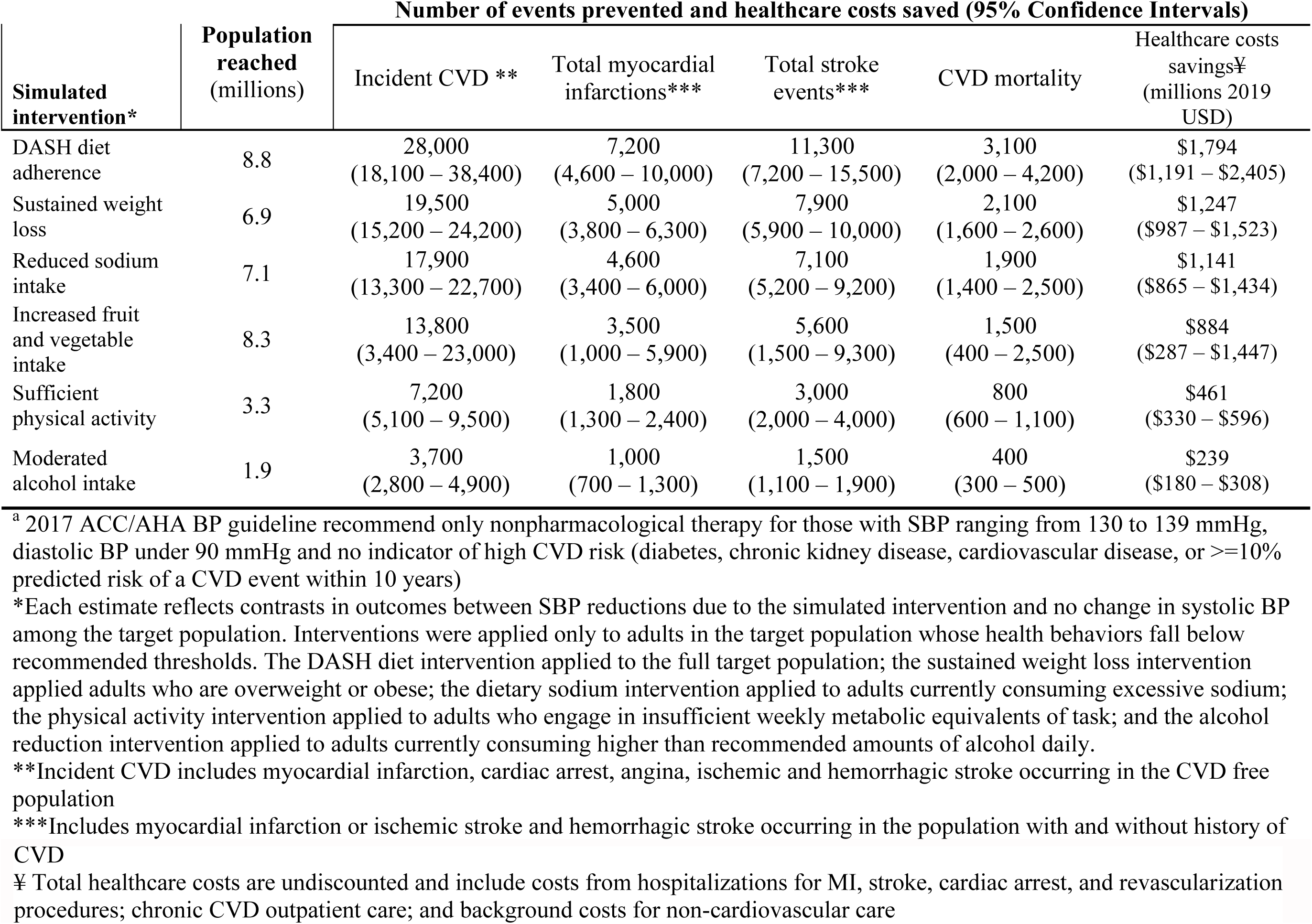
Projected impact of evidence-based nonpharmacological therapy in adults with stage 1 hypertension by 2017 ACC/AHA BP guideline^a^, 2018-2027.

| Simulated intervention* | Population reached (millions) | Number of events prevented and healthcare costs saved (95% Confidence Intervals) |  |  |  |  |
| --- | --- | --- | --- | --- | --- | --- |
|  |  | Incident CVD ** | Total myocardial infarctions*** | Total stroke events*** | CVD mortality | Healthcare costs savings¥ (millions 2019 USD) |
| DASH diet adherence | 8.8 | 28,000<br>(18,100 – 38,400) | 7,200<br>(4,600 – 10,000) | 11,300<br>(7,200 – 15,500) | 3,100<br>(2,000 – 4,200) | \$1,794<br>(\$1,191 – \$2,405) |
| Sustained weight loss | 6.9 | 19,500<br>(15,200 – 24,200) | 5,000<br>(3,800 – 6,300) | 7,900<br>(5,900 – 10,000) | 2,100<br>(1,600 – 2,600) | \$1,247<br>(\$987 – \$1,523) |
| Reduced sodium intake | 7.1 | 17,900<br>(13,300 – 22,700) | 4,600<br>(3,400 – 6,000) | 7,100<br>(5,200 – 9,200) | 1,900<br>(1,400 – 2,500) | \$1,141<br>(\$865 – \$1,434) |
| Increased fruit and vegetable intake | 8.3 | 13,800<br>(3,400 – 23,000) | 3,500<br>(1,000 – 5,900) | 5,600<br>(1,500 – 9,300) | 1,500<br>(400 – 2,500) | \$884<br>(\$287 – \$1,447) |
| Sufficient physical activity | 3.3 | 7,200<br>(5,100 – 9,500) | 1,800<br>(1,300 – 2,400) | 3,000<br>(2,000 – 4,000) | 800<br>(600 – 1,100) | \$461<br>(\$330 – \$596) |
| Moderated alcohol intake | 1.9 | 3,700<br>(2,800 – 4,900) | 1,000<br>(700 – 1,300) | 1,500<br>(1,100 – 1,900) | 400<br>(300 – 500) | \$239<br>(\$180 – \$308) |

<sup>a</sup> 2017 ACC/AHA BP guideline recommend only nonpharmacological therapy for those with SBP ranging from 130 to 139 mmHg, diastolic BP under 90 mmHg and no indicator of high CVD risk (diabetes, chronic kidney disease, cardiovascular disease, or $\geq 10\%$ predicted risk of a CVD event within 10 years)
\*Each estimate reflects contrasts in outcomes between SBP reductions due to the simulated intervention and no change in systolic BP among the target population. Interventions were applied only to adults in the target population whose health behaviors fall below recommended thresholds. The DASH diet intervention applied to the full target population; the sustained weight loss intervention applied adults who are overweight or obese; the dietary sodium intervention applied to adults currently consuming excessive sodium; the physical activity intervention applied to adults who engage in insufficient weekly metabolic equivalents of task; and the alcohol reduction intervention applied to adults currently consuming higher than recommended amounts of alcohol daily.
\*\*Incident CVD includes myocardial infarction, cardiac arrest, angina, ischemic and hemorrhagic stroke occurring in the CVD free population
\*\*\*Includes myocardial infarction or ischemic stroke and hemorrhagic stroke occurring in the population with and without history of

## Discussion

Nearly 9 million US adults aged 35-64 years with stage 1 systolic hypertension would be recommended nonpharmacological therapy by the 2017 ACC/AHA BP guideline because of younger age and lower CVD risk. We projected that controlling SBP to <130 mmHg could prevent over 26,000 CVD events compared with the status quo. Universal adoption of the DASH diet was projected to prevent 28,000 CVD events over 10 years. Sustained weight loss, sodium reduction, and increased fruit and vegetable intake could also offer significant population benefits. Finally, relying solely on behavioral counseling at healthcare encounters might fail to reach the half of men and one-quarter of women with stage 1 systolic hypertension at low CVD risk who do not regularly engage with the healthcare system.

As the JNC7 BP guideline explicitly stated prehypertension was not a disease category^1^, the lower BP threshold to define hypertension in the 2017 ACC/AHA BP guideline is an example of medicalization that effectively created a new medical diagnosis for those previously considered disease-free ^38^. A new diagnosis of hypertension may change incentives in physician-patient interactions. However, many providers have limited training in in effective health counseling, let alone in tailoring recommendations for diverse patient populations ^39^. A common critique of medicalization is that racial and ethnic minorities and people of lower socioeconomic status (SES) face more barriers that prevent them from fulfilling these ‘prescribed’ health behavior changes. Furthermore, the population most in need of nonpharmacological BP control is often uninsured, and does not regularly access healthcare for counseling opportunities to occur^40^.

Policies that improve the food system and neighborhood conditions may facilitate recommended behavior changes irrespective of whether a person regularly interacts with a healthcare provider. Though salt alternatives can provide a more favorable ratio of dietary potassium to sodium among those who cook at home^41^ population benefits may be minimal among groups reliant on processed foods without commitment from stakeholders in restaurant and food manufacturing industries^42^. The US Food and Drug Administration has recently announced voluntary sodium reduction guidelines for commercially produced and packaged foods, involving gradually reducing maximum sodium content to allow consumer tastes to adapt and focusing on per-package versus per-serving sodium content^43^. Similar industry-oriented policies in the United Kingdom were associated with modest annual reductions in sodium levels detected in population-representative urine samples between 2003 and 2007^44^.

Community members are more likely to be more physically active if they reside in economically invested areas that are well-connected by sidewalks, with adequate street lighting and lower levels of crime^45,46^. Amenities within walking distance that have been associated with lower prevalence of physical inactivity and obesity include destinations for errands, green spaces such as parks, outdoor features such as beaches and trails, and a public transit system that is proximate to employment^47,48^. Racial and ethnic minorities as well as low socioeconomic status people are concentrated in areas with more barriers to walking, fewer free physical activity facilities, and limited available healthy food choices^46,49^. Marginalized people also often report having less time and resources to engage in recommended health behaviors^50^. Multilevel public health strategies that restructure the disadvantaged environmental context for vulnerable groups are vital to redress CVD disparities. These public health approaches can synergistically achieve a broader population reach than the individual behavior change interventions that provided inputs for our analysis. Quantifying the cost of such population-level interventions would enable estimation of cost-effectiveness.

## Limitations

Our study integrated several nationally representative data sources into a platform that simulated the future impact of potential population-level interventions on blood pressure control. However, NHANES only provides participant self-report of health behaviors such as diet and physical activity, which may be subject to reporting bias. We modeled 50% and 100% uptake of each nonpharmacological intervention and assumed sustained SBP reduction over 10 years, which may be difficult to achieve in practice. Our results may therefore represent projections of the upper limit of population benefits with independent interventions. We used meta-analysis and trial results to simulate SBP reductions with nonpharmacologic therapy, where information on long-term adherence was unavailable. Though our methods could only simulate the independent impact of each intervention, nonpharmacological SBP management realistically necessitates a combination of health behaviors: e.g., weight reduction via sufficient exercise in conjunction with dietary modification. Our analysis only included individuals with stage 1 hypertension according to the 2017 ACC/AHA BP guideline; however, if implemented population-wide, nonpharmacological therapy could have even greater benefit by preventing CVD events among individuals with normal and elevated BP as well as patients already on antihypertensive therapy.

## Conclusions

We used a computer simulation model to project the potential health and economic benefits of implementing evidence-based nonpharmacological therapy among nearly 9 million US adults with stage 1 systolic hypertension. Each intervention has the potential to prevent between 3,700 and 28,000 CVD events. Full adherence of the DASH diet could save up to 3,100 lives and advert upwards of $1.7 billion in healthcare costs over 10 years. However, as only two-thirds of these individuals regularly interact with the healthcare system, achieving the full benefit via recommended provider counseling may remain elusive. Even among those in care, marginalized people systematically lack the individual- or community-level resources needed to modify their health behavior. For maximal population benefit, food system policies that improve accessibility to nutrient-rich food as well as lower sodium choices will facilitate large-scale adoption. Policymakers, industry stakeholders, and medical professionals need to collaborate on strategies to manage blood pressure among this large group of adults vulnerable to long-term CVD risk. Broad public health interventions on the local and federal infrastructure would likely result in substantial benefits not only for adults with Stage 1 hypertension, but for the U.S. population at large.

## Data Availability

National Health and Nutrition Examination Survey is publicly available.

## Non-standard Abbreviations and Acronyms

ACC/AHA: American College of Cardiology and American Heart Association (ACC/AHA)
SBP: systolic blood pressure
DBP: diastolic blood pressure
DASH: Dietary Approaches to Stop Hypertension
NHANES: National Health and Nutrition Examination Survey

